# Door-in-Door-Out Times at Referring Hospitals and Outcomes from Hemorrhagic Stroke

**DOI:** 10.1101/2024.04.26.24306465

**Authors:** Regina Royan, Iyanuoluwa Ayodele, Brian Stamm, Brooke Alhanti, Kevin N. Sheth, Peter Pruitt, Brian Mac Grory, William J. Meurer, Shyam Prabhakaran

## Abstract

**Background:** Guidelines recommend DIDO (Door-In-Door-Out) time ≤120 minutes at the transferring emergency department (ED); however, it is unknown whether inter-hospital transfer times are related to clinical outcomes.

**Methods:** Retrospective, observational cohort study using US registry data from GWTG-Stroke participating hospitals. Patients age ≥18 years with intracerebral hemorrhage (ICH) or subarachnoid hemorrhage (SAH) from January 1, 2019 to July 31, 2022 who were transferred from the ED to a GWTG-participating receiving hospital. Outcomes included discharge modified Rankin Score (mRS) 0-3 vs 4-6; ability to ambulate independently at discharge; and in-hospital mortality at the receiving hospital.

**Results:** In total, 19,708 ICH patients were included, with median age 68.0 years (IQR 57.0-78.0), 46.2% female, 65.2% White, 16.8% Black, and 8.5% Hispanic. 7,757 SAH patients were included, with median age 59.0 years (IQR 49.0-69.0), 62.7% female, 62.0% White, 14.6% Black, and 11.9% Hispanic. For ICH patients, increasing DIDO time was associated with greater odds of mRS 0-3 vs 4-6 at discharge in the unadjusted analyses (DIDO 91-180 mins, OR 1.15 [1.04-1.27]; 181-270 mins, OR 1.51 [1.33, 1.71]; >270 mins, OR 1.83 [1.58, 2.11]; vs DIDO ≤ 90 mins as reference; P<.0001), but these associations became statistically non-significant in the adjusted analyses. Similar results were seen for mRS at discharge in SAH patients. In both ICH and SAH patients, longer DIDO times were associated with greater odds of independent ambulation at discharge and lower odds of in-hospital mortality.

**Conclusion:** DIDO times were inversely related to in-hospital mortality, ability to ambulate independently at discharge, but not discharge mRS for patients with ICH and SAH. These findings may suggest that a longer period of stabilization in the initial ED may be associated with better outcomes from hemorrhagic stroke and that current interhospital transfer protocols currently expedite transfer of the sickest patients. Prospective studies are needed to balance ED stabilization with arrival at a definitive destination in patients with hemorrhagic stroke.

**KEY POINTS:** *Question:* Is Door-In-Door Out time at a transferring hospital associated with global disability at hospital discharge (modified Rankin Score (mRS))for patients with intracranial hemorrhage?

*Findings:* In this cohort study using a large nationwide quality improvement database, increasing DIDO time was associated with greater odds of mRS 0-3 vs 4-6 at discharge in the unadjusted analyses for both ICH and SAH patients, but these associations became statistically non-significant in the adjusted analyses. In both ICH and SAH patients, longer DIDO times were associated with greater odds of independent ambulation at discharge and lower odds of in-hospital mortality in both the unadjusted and adjusted analyses.

*Meaning:* These findings may suggest that a longer period of stabilization in the initial emergency department (ED) may be associated with better outcomes from hemorrhagic stroke and that current interhospital transfer protocols tend to expedite transfer of the sickest patients. Prospective studies are needed to determine whether early or delayed transport with ED stabilization is optimal for care of patients with hemorrhagic stroke.

## INTRODUCTION

While standardized performance metrics for acute emergency care of acute ischemic stroke are now ubiquitous, such measures are less prevalent in the treatment of hemorrhagic stroke.^1^ Yet, the management of hemorrhagic stroke involves expeditious blood pressure control,^2–4^ reversal of coagulopathy,^2,5^ and medical stabilization.^6^ The INTERACT3 trial showed that early bundled treatment of elevated blood pressure, fever, hyperglycemia, and coagulopathy reversal within several hours of the onset of symptoms resulted in improved functional outcome for patients with acute intracerebral hemorrhage (ICH).^7^ A recent study from the Get With the Guidelines (GWTG)-Stroke registry demonstrated an association between earlier anticoagulation reversal and improved survival from ICH.^8^ Additionally, a 2024 systematic review from the Blood Pressure in Acute Stroke Collaboration showed a clear time relation between early vs late blood pressure control in ICH patients with improvement in functional outcomes and a lower risk of hematoma expansion.^4^ Ongoing efforts including the recent consensus statement for “Code ICH” emphasize the importance of early bundled care for ICH and adherence to time-based metrics,^9^ which could change the landscape of care for hemorrhagic stroke.

Door-in-door-out (DIDO) time, defined as the arrival to discharge time in the initial ED prior to transfer for higher level of care,^10^ has emerged as a quality metric in the management of acute stroke. In a study using the national GWTG-Stroke registry, the median DIDO time for patients with hemorrhagic stroke was 178 minutes (IQR, 119-275 minutes),^15^ significantly exceeding recommended benchmarks. Previous literature found that prolonged ED boarding times were independently associated with poor outcomes in acute stroke patients awaiting transfer to the Neuro-ICU or stroke inpatient units.^16,17^ However, the relationship between DIDO time and functional outcomes in hemorrhagic stroke has not been previously evaluated.

We used the GWTG-Stroke registry to examine the relationship between DIDO time and modified Rankin scale (mRS) score at discharge in patients transferred for management of hemorrhagic stroke. We hypothesized that shorter DIDO times would be associated with more favorable outcomes from hemorrhagic stroke.

## METHODS

Cases were obtained from the American Heart Association GWTG-Stroke registry, a voluntary, national quality improvement database which is representative of the United States Medicare ischemic stroke population.^18^ Each participating hospital received either human research approval to enroll cases without individual patient consent under the common rule, or a waiver of authorization and exemption from subsequent review by their institutional review board. The Duke Clinical Research Institute serves as the data analysis center and has an agreement to analyze the aggregate limited data for research purposes. The Institutional Review Board at Duke University Health approved this study. IQVIA (Parsippany, New Jersey) serves as the data collection and coordination center. This study follows STROBE reporting guidelines for observational studies.^19^

### Study Population

The study population was stratified by hemorrhagic stroke type: ICH and subarachnoid hemorrhage (SAH). Patients with ICH and/or SAH who were transferred from another hospital and admitted to GWTG-Stroke hospitals from Jan 1, 2019 to Jul 31, 2022, age ≥18 years with non-missing sex, and from sites with <25% missing data on the medical history panel were included. Patients with post-thrombolytic hemorrhagic stroke, DIDO time >24 hours, missing DIDO time, patients who left against medical advice, discharge information missing or not documented, and National Institute of Health Stroke Scale (NIHSS) information missing were excluded.

### Outcomes

The pre-specified primary outcome was ordinal mRS at discharge (0, 1, 2, 3, 4, 5-6 scale); with mRS 5 and 6 combined while modeling in order to increase likelihood of meeting the proportional odds assumption. Secondary outcomes included the following binary variables: mRS at discharge (0-3 vs 4-6); ambulate independently at discharge (among those patients who were ambulatory prior to admission); in-hospital mortality; and in-hospital mortality or discharged to hospice. An additional secondary outcome was the utility-weighted mRS at discharge (UW-mRS).^20^

### Pre-specified covariates

The primary exposure was DIDO time, defined as referring hospital arrival time to discharge time prior to transfer.

The following factors were adjusted for in the models. Demographic factors included age, sex, and insurance status. Medical history variables included atrial fibrillation/flutter, previous stroke/transient ischemic attack (TIA), coronary artery disease (CAD)/prior myocardial infarction (MI), prior heart failure (HF), carotid stenosis, diabetes, peripheral vascular disease, hypertension, dyslipidemia, prosthetic heart valve, smoker, and renal insufficiency. Admission characteristics included: NIHSS score, antiplatelet or anticoagulant use, systolic blood pressure (SBP), and international normalized ratio (INR). Receiving hospital characteristics included region, rural location, teaching hospital, number of beds, annual hemorrhagic stroke volume, and stroke center status.

### Missing Data

Model covariates with missing data were imputed before entering into models following previous GWTG-stroke studies.^8,21^ Observations with >25% missing were excluded and missing medical history values and medications prior to admission were imputed to “No”. Insurance status for patients 65 years or older was imputed to “Medicare” and all other patients were imputed to “Private / VA / Champus / Other Insurance”. Teaching hospital status and rural hospital location were imputed to “No”. Hospital size was imputed to the conditional median hospital size for teaching sites (430) and for non-teaching sites (210). Patients with in-hospital mortality with missing mRS data were imputed to 6, otherwise multiple imputation methods were used with 20 datasets for variables with ≤25% missing data.

### Statistical Analysis

Baseline patient characteristics were reported using proportions for categorical variables and median with first and third quartiles for continuous variables. Procedural outcomes overall and by DIDO time (≤90mins, 91-180mins, 181-270 mins, >270 mins) were also described for ICH patients.

The association between the DIDO time and clinical outcomes for patients was assessed, stratified by ICH and SAH groups. Multivariable regression models with generalized estimating equations (GEE) and independent working correlation structure were used in models to account for within and across hospital variability. An ordinal logistic model was planned for the primary ordinal outcome with a report of the unadjusted and adjusted proportional odds ratios. However, the proportional odds assumption was violated; therefore this was omitted from the included analysis. Unadjusted comparison of groups was performed using the Kruskal Wallis test.

Logistic regression was used to report unadjusted and adjusted odds ratios for the secondary outcomes of mRS at discharge (0-3 vs 4-6); independent ambulation at discharge (among those patients who were ambulatory prior to admission); in-hospital mortality; and in-hospital mortality or discharge to hospice. Beta regression was used for the secondary weighted outcome, with a random effect to account for within and across hospital variability with hospital-specific random intercepts. Unadjusted and adjusted odds ratios were reported for UW-mRS. UW-mRS scores were assigned utility values of: mRS 0 – 1; mRS 1 – 0.91; mRS 2 – 0.76; mRS 3 – 0.65; mRS 4 – 0.33; mRS 5 – 0; mRS 6 – 0.^22^ To account for the non-inclusivity limitation of 0 or 1 in the Beta regression model, the UW-mRS scores were transformed using N-1(y*(N-1)+0.5), where y is the UW-mRS score and N is the total study population.^23^ Unadjusted and adjusted odds and proportional odds ratios were also reported among ICH and SAH patients with last-known well (LKW) to outside hospital arrival time ≤ 120 mins.

Supplemental analysis included examination of temporal trends in DIDO time by referring admission quarters using the Cochran-Armitage trend test, as well as a comparison of baseline patient characteristics for patients with DIDO ≤ 24 hours vs. those with missing DIDO information in the ICH and SAH study populations. All statistical tests were two-sided and utilized the nominal type I error rate ɑ=0.05 or absolute standardized difference of <10% to be considered significant. Analysis was performed at the Duke Clinical Research Institute using SAS software version 9.4 (SAS Institute, Cary, North Carolina) on the American Heart Association Precision Medicine Platform (https://precision.heart.org).

## RESULTS

In total, 19,708 ICH patients were included (Supplemental Figure 1). The overall ICH population had a median age of 68.0 years (IQR 57.0-78.0), was 46.2% female, 65.2% White, 16.8% Black, and 8.5% Hispanic. Median NIHSS score was 6.0 (IQR 2.0-18.0). Receiving hospitals were most likely to be in the South (39.3%), and the majority were comprehensive stroke centers (54.4%). Median DIDO time was 155.0 minutes (IQR 101.0-243.0), and median mRS score at discharge was 4.0 for ICH (IQR 3.0-5.0) (Table 1).

**Table 1.** Patient and hospital characteristics for patients with intracerebral hemorrhage (ICH) and subarachnoid hemorrhage (SAH)

|  | ICH<br>(N=19,708) | SAH<br>(N=7,757) |
| --- | --- | --- |
| **Demographics** |  |  |
| Age, years | 68.0 (57.0-78.0) | 59.0 (49.0-69.0) |
| Female | 9,106 (46.2%) | 4,860 (62.7%) |
| Race-Ethnicity |  |  |
| White | 12,839 (65.2%) | 4,809 (62.0%) |
| Black | 3,309 (16.8%) | 1,134 (14.6%) |
| Hispanic (any race) | 1,678 (8.5%) | 922 (11.9%) |
| Asian | 778 (3.9%) | 354 (4.6%) |
| Other (includes UTD) | 1,101 (5.6%) | 538 (6.9%) |
| Insurance |  |  |
| Private/VA/Champus/Other Insurance | 6,814 (36.1%) | 3,609 (48.6%) |
| Medicaid | 2,979 (15.8%) | 1,231 (16.6%) |
| Medicare | 7,641 (40.5%) | 1,880 (25.3%) |
| Other (self, none, ND) | 1,416 (7.5%) | 704 (9.5%) |
| **Medical History** |  |  |
| Atrial fibrillation/flutter | 3,229 (16.4%) | 458 (5.9%) |
| Previous Stroke/TIA | 4,532 (23.0%) | 744 (9.6%) |
| Coronary Artery Disease/Myocardial Infarction | 3,341 (17.0%) | 682 (8.8%) |
| Heart Failure | 1,580 (8.0%) | 288 (3.7%) |
| Carotid stenosis | 373 (1.9%) | 98 (1.3%) |
| Diabetes | 5,234 (26.6%) | 1,215 (15.7%) |
| Peripheral vascular disease | 541 (2.7%) | 165 (2.1%) |
| Hypertension | 14,841 (75.3%) | 4,384 (56.5%) |
| Dyslipidemia | 8,153 (41.4%) | 2,386 (30.8%) |
| Prosthetic heart valve | 282 (1.4%) | 56 (0.7%) |
| Smoker | 3,128 (15.9%) | 1,982 (25.6%) |
| Renal insufficiency | 2,097 (10.6%) | 391 (5.0%) |
| **Clinical Characteristics** |  |  |
| SBP, mmHg | 152.0 (135.0-175.0) | 146.0 (130.0-166.0) |
| INR | 1.1 (1.0-1.2) | 1.0 (1.0-1.1) |
| NIHSS score, (median) | 6.0 (2.0-18.0) | 1.0 (0.0-12.0) |
| NIHSS group |  |  |
| 0-4 | 8,368 (42.5%) | 5,028 (64.8%) |
| 5-9 | 3,102 (15.7%) | 585 (7.5%) |
| 10-14 | 1,958 (9.9%) | 365 (4.7%) |
| 15-20 | 2,285 (11.6%) | 486 (6.3%) |
| >20 | 3,995 (20.3%) | 1,293 (16.7%) |
| Door-in-door-out time, min | 155.0 (101.0-243.0) | 179.0 (120.0-273.0) |
| **Medications prior to admission** |  |  |
| Antiplatelets | 6,245 (31.7%) | 1,552 (20.0%) |
| Anticoagulants | 3,436 (17.4%) | 468 (6.0%) |
| **Clinical outcomes** |  |  |
| Modified Rankin score at discharge |  |  |
| 0 | 846 (5.7%) | 1,077 (18.5%) |
| 1 | 1,259 (8.5%) | 940 (16.1%) |
| 2 | 1,079 (7.3%) | 539 (9.2%) |
| 3 | 1,671 (11.3%) | 599 (10.3%) |
| 4 | 4,468 (30.3%) | 1,043 (17.9%) |
| 5 | 2,604 (17.6%) | 631 (10.8%) |
| 6 | 2,831 (19.2%) | 1,007 (17.3%) |
| mRS, Median (IQR) | 4.0 (3.0-5.0) | 3.0 (1.0-5.0) |
| mRS, Mean (SD) | 3.8 (1.7) | 2.9 (2.1) |
| UW-mRS | 0.3 (0.0-0.7) | 0.7 (0.0-0.9) |
| Ambulatory status at discharge |  |  |
| Able to ambulate independently | 6,163 (32.3%) | 4,056 (54.2%) |
| With assistance from person | 6,308 (33.1%) | 1,533 (20.5%) |
| Unable to ambulate | 3,775 (19.8%) | 893 (11.9%) |
| In-hospital death | 2,831 (14.8%) | 1,007 (13.4%) |
| Discharge disposition |  |  |
| Home | 6,157 (42.7%) | 4,297 (65.5%) |
| Acute care facility | 325 (2.3%) | 168 (2.6%) |
| Skilled Nursing facility | 3,138 (21.7%) | 586 (8.9%) |
| Long term care hospital | 540 (3.7%) | 211 (3.2%) |
| Hospice | 1,438 (10.0%) | 295 (4.5%) |
| In-hospital mortality | 2,831 (19.6%) | 1,007 (15.3%) |
| **Hospital Characteristics** |  |  |
| Region |  |  |
| Northeast | 4,345 (22.0%) | 1,544 (19.9%) |
| Midwest | 4,683 (23.8%) | 1,740 (22.4%) |
| South | 7,743 (39.3%) | 3,185 (41.1%) |
| West | 2,937 (14.9%) | 1,288 (16.6%) |
| Rural Location | 243 (1.2%) | 24 (0.3%) |
| No. of beds | 516 (381-764) | 528 (384-773) |
| Academic/Teaching Hospital | 17,583 (91.2%) | 6,905 (91.7%) |
| Primary Stroke Center status |  |  |
| Comprehensive Stroke Center (CSC) | 10,718 (54.4%) | 4,654 (60.0%) |
| Primary Stroke Center (PSC) | 6,386 (32.4%) | 2,199 (28.3%) |
| Neither CSC or PSC | 2,604 (13.2%) | 904 (11.7%) |
| Annual ICH stroke | 87.8 (60.6-138.3) | 33.9 (23.2-59.0) |

7,757 SAH patients were included. The overall SAH population had a median age of 59.0 years (IQR 49.0-69.0), was 62.7% female, 62.0% White, 14.6% Black, and 11.9% Hispanic. Median NIHSS score was 1.0 (IQR 0.0-12.0). Receiving hospitals were most likely to be in the South (41.1%), and the majority were comprehensive stroke centers (60.0%). Median DIDO time was 179.0 minutes (IQR 120.0-273.0), and median mRS score at discharge was 3.0 (IQR 1.0-5.0); (Table 1). More severe strokes with higher NIHSS scores had greater proportions of patients with DIDO times of <=90 mins and 91-180 mins (Supplemental Tables 1 and 2)

Surgical treatments for ICH patients differed between the DIDO time tertiles, namely the proportions of external ventricular drain (EVD) placement and conventional craniotomy / evacuation of clot under direct vision had greater proportions of patients transferred in ≤ 90 minutes and 91-180 minutes (Table 2).

**Table 2.**
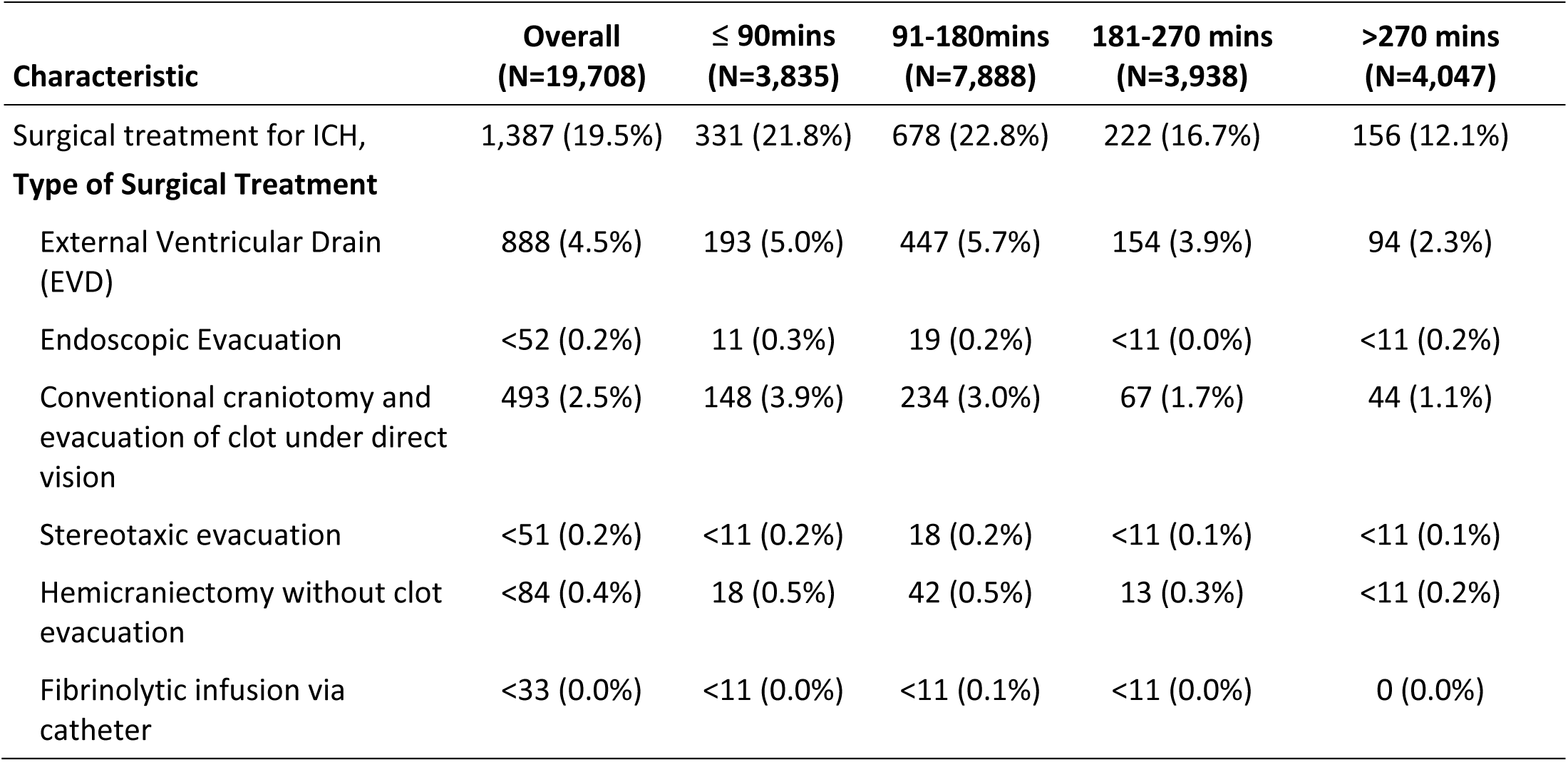
Procedural outcomes of ICH patients, stratified by DIDO time.

For ICH patients in the unadjusted analysis, increasing DIDO time (by tertile) was associated with greater odds of mRS 0-3 vs 4-6 at discharge: DIDO 91-180 mins, OR 1.15 (95% CI 1.04, 1.27); DIDO 181-270 mins, OR 1.51 (95% CI 1.33, 1.71); and DIDO >270mins, OR 1.83 (95% CI 1.58, 2.11), compared to reference of DIDO ≤90mins. After adjustment for covariates in the models, these associations became statistically non-significant. Results were similar for the outcome of utility-weighted mRS at discharge. For ICH patients in the unadjusted analysis, increasing DIDO time was associated with greater odds of independent ambulation at discharge: DIDO 91-180 mins, OR 1.23 (1.11, 1.36); DIDO 181-270 mins, OR 1.76 (1.54, 2.00); and DIDO >270mins, OR 2.16 (1.88, 2.48), compared to reference of DIDO ≤90mins. After adjustment, the effect sizes of these associations were reduced, but the results remained statistically significant (adjusted P=0.043). Similarly, in the unadjusted analysis, increasing DIDO time was associated with lower odds of in-hospital mortality: DIDO 91-180 mins, OR 1.00 (0.86, 1.16); DIDO 181-270 mins, OR 0.67 (0.57, 0.80); and DIDO >270mins, OR 0.52 (0.43, 0.62), compared to reference of DIDO ≤90mins. After adjustment, the effect sizes of these associations were reduced, but the results remained statistically significant (adjusted P=0.026). Similar results were seen for the outcome of in-hospital mortality or discharge to hospice, but after adjustment, the associations became statistically non-significant (Table 3). Results were overall similar for a subgroup analysis of ICH patients with LKW to outside hospital arrival times ≤ 120 mins (Supplemental Table 3). Unadjusted distributions were graphically depicted using stacked bar charts (Grotta bars; Supplemental Figures 2a and 2b).

**Table 3.**
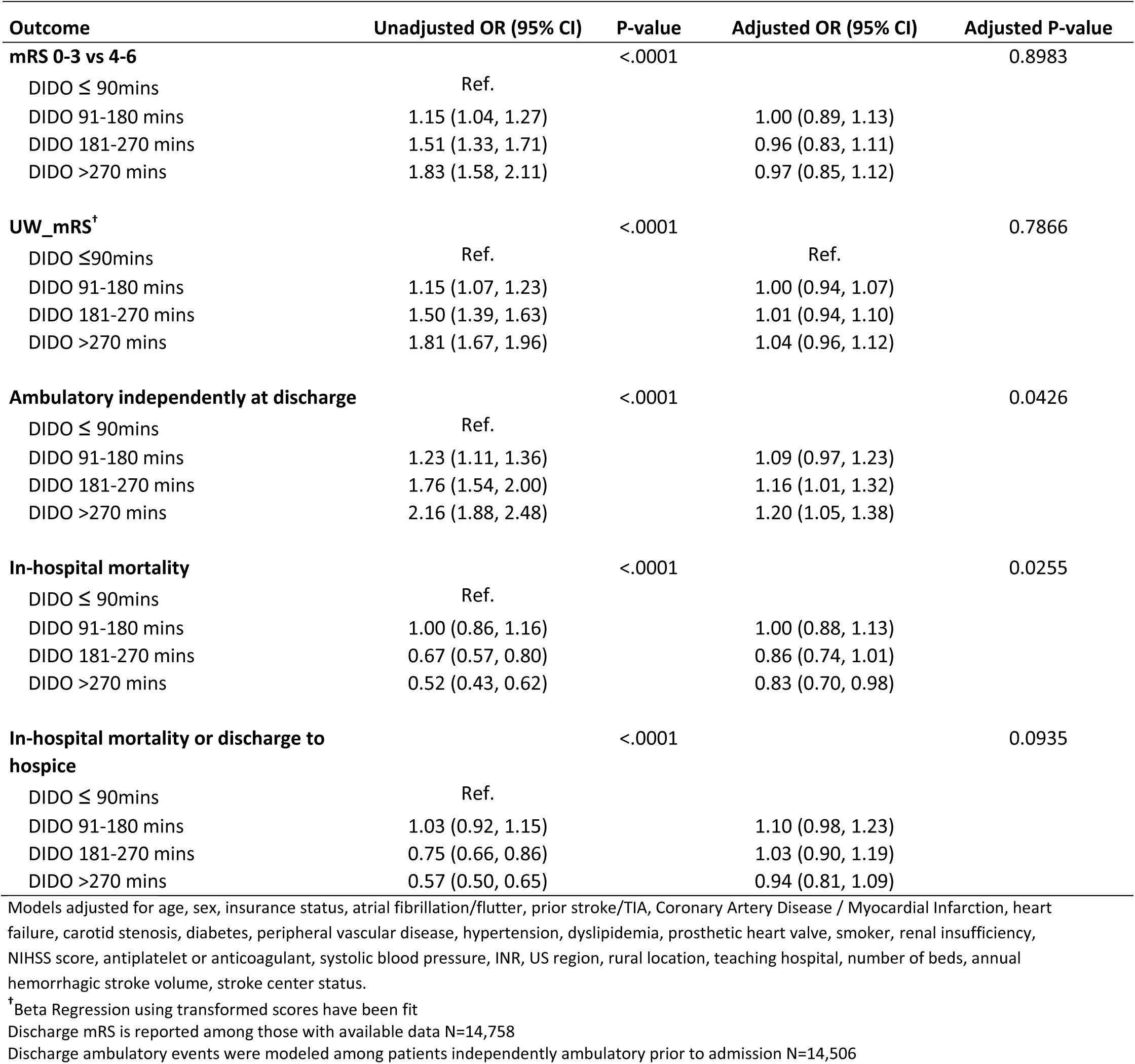
Association of DIDO time with clinical outcomes for ICH patients.

| Outcome | Unadjusted OR (95% CI) | P-value | Adjusted OR (95% CI) | Adjusted P-value |
| --- | --- | --- | --- | --- |
| **mRS 0-3 vs 4-6** |  | <.0001 |  | 0.8983 |
| DIDO ≤ 90mins | Ref. |  |  |  |
| DIDO 91-180 mins | 1.15 (1.04, 1.27) |  | 1.00 (0.89, 1.13) |  |
| DIDO 181-270 mins | 1.51 (1.33, 1.71) |  | 0.96 (0.83, 1.11) |  |
| DIDO >270 mins | 1.83 (1.58, 2.11) |  | 0.97 (0.85, 1.12) |  |
| **UW_mRS<sup>†</sup>** |  | <.0001 |  | 0.7866 |
| DIDO ≤90mins | Ref. |  | Ref. |  |
| DIDO 91-180 mins | 1.15 (1.07, 1.23) |  | 1.00 (0.94, 1.07) |  |
| DIDO 181-270 mins | 1.50 (1.39, 1.63) |  | 1.01 (0.94, 1.10) |  |
| DIDO >270 mins | 1.81 (1.67, 1.96) |  | 1.04 (0.96, 1.12) |  |
| **Ambulatory independently at discharge** |  | <.0001 |  | 0.0426 |
| DIDO ≤ 90mins | Ref. |  |  |  |
| DIDO 91-180 mins | 1.23 (1.11, 1.36) |  | 1.09 (0.97, 1.23) |  |
| DIDO 181-270 mins | 1.76 (1.54, 2.00) |  | 1.16 (1.01, 1.32) |  |
| DIDO >270 mins | 2.16 (1.88, 2.48) |  | 1.20 (1.05, 1.38) |  |
| **In-hospital mortality** |  | <.0001 |  | 0.0255 |
| DIDO ≤ 90mins | Ref. |  |  |  |
| DIDO 91-180 mins | 1.00 (0.86, 1.16) |  | 1.00 (0.88, 1.13) |  |
| DIDO 181-270 mins | 0.67 (0.57, 0.80) |  | 0.86 (0.74, 1.01) |  |
| DIDO >270 mins | 0.52 (0.43, 0.62) |  | 0.83 (0.70, 0.98) |  |
| **In-hospital mortality or discharge to hospice** |  | <.0001 |  | 0.0935 |
| DIDO ≤ 90mins | Ref. |  |  |  |
| DIDO 91-180 mins | 1.03 (0.92, 1.15) |  | 1.10 (0.98, 1.23) |  |
| DIDO 181-270 mins | 0.75 (0.66, 0.86) |  | 1.03 (0.90, 1.19) |  |
| DIDO >270 mins | 0.57 (0.50, 0.65) |  | 0.94 (0.81, 1.09) |  |

Models adjusted for age, sex, insurance status, atrial fibrillation/flutter, prior stroke/TIA, Coronary Artery Disease / Myocardial Infarction, heart failure, carotid stenosis, diabetes, peripheral vascular disease, hypertension, dyslipidemia, prosthetic heart valve, smoker, renal insufficiency, NIHSS score, antiplatelet or anticoagulant, systolic blood pressure, INR, US region, rural location, teaching hospital, number of beds, annual hemorrhagic stroke volume, stroke center status.
<sup>†</sup> Beta Regression using transformed scores have been fit
Discharge mRS is reported among those with available data N=14,758
Discharge ambulatory events were modeled among patients independently ambulatory prior to admission N=14,506

For SAH patients in the unadjusted analysis, increasing DIDO time (by tertile) was associated with greater odds of mRS 0-3 vs 4-6 at discharge: DIDO 91-180 mins, OR 1.28 (1.04, 1.57); DIDO 181-270 mins, OR 1.49 (1.20, 1.86); and DIDO >270mins, OR 1.82 (1.47, 2.27), compared to reference of DIDO ≤90mins. After adjustment for covariates in the models, these associations became statistically non-significant. Results were similar for the outcome of utility-weighted mRS at discharge in the unadjusted analysis and remained statistically significant in the adjusted analysis (adjusted P-value 0.0009). In the unadjusted analysis, increasing DIDO time was associated with greater odds of independent ambulation at discharge: DIDO 91-180 mins, OR 1.27 (1.04, 1.55); DIDO 181-270 mins, OR 1.50 (1.20, 1.88); and DIDO >270mins, OR 1.90 (1.54, 2.33), compared to reference of DIDO ≤90mins. After adjustment, the effect sizes of these associations were reduced, but the results remained statistically significant (adjusted P=0.008). Similarly, in the unadjusted analysis, increasing DIDO time was associated with lower odds of in-hospital mortality: DIDO 91-180 mins, OR 0.81 (0.67, 0.98); DIDO 181-270 mins, 0.62 (0.50, 0.78); and DIDO >270mins, OR 0.43 (0.34, 0.54), compared to reference of DIDO ≤90mins. After adjustment, the effect sizes of these associations were reduced, but the results remained statistically significant (adjusted P=0.003). Similar results were seen for the outcome of in-hospital mortality or discharge to hospice (Table 4). Subgroup analysis of SAH patients with LKW to outside hospital arrival times ≤ 120 mins were similar (Supplemental Table 4).

**Table 4.** Association of DIDO time with clinical outcomes for SAH patients.

| Outcome | Unadjusted OR (95% CI) | P-value | Adjusted OR (95% CI) | Adjusted P-value |
| --- | --- | --- | --- | --- |
| **mRS 0-3 vs 4-6** |  | <.0001 |  | 0.1348 |
| DIDO ≤ 90mins | Ref. |  |  |  |
| DIDO 91-180 mins | 1.28 (1.04, 1.57) |  | 1.14 (0.92, 1.42) |  |
| DIDO 181-270 mins | 1.49 (1.20, 1.86) |  | 1.24 (0.98, 1.56) |  |
| DIDO >270 mins | 1.82 (1.47, 2.27) |  | 1.30 (1.03, 1.64) |  |
| **UW_mRS<sup>†</sup>** |  | <.0001 |  | 0.0009 |
| DIDO ≤ 90mins | Ref. |  | Ref. |  |
| DIDO 91-180 mins | 1.29 (1.13, 1.47) |  | 1.10 (0.96, 1.27) |  |
| DIDO 181-270 mins | 1.52 (1.31, 1.76) |  | 1.21 (1.04, 1.40) |  |
| DIDO >270 mins | 1.90 (1.64, 2.20) |  | 1.32 (1.14, 1.53) |  |
| **Ambulatory independently at discharge** |  | <.0001 |  | 0.0082 |
| DIDO ≤ 90mins | Ref. |  |  |  |
| DIDO 91-180 mins | 1.27 (1.04, 1.55) |  | 1.07 (0.87, 1.32) |  |
| DIDO 181-270 mins | 1.50 (1.20, 1.88) |  | 1.20 (0.96, 1.50) |  |
| DIDO >270 mins | 1.90 (1.54, 2.33) |  | 1.39 (1.11, 1.74) |  |
| **In-hospital mortality** |  | <.0001 |  | 0.0028 |
| DIDO ≤ 90mins | Ref. |  |  |  |
| DIDO 91-180 mins | 0.81 (0.67, 0.98) |  | 0.84 (0.66, 1.06) |  |
| DIDO 181-270 mins | 0.62 (0.50, 0.78) |  | 0.74 (0.57, 0.97) |  |
| DIDO >270 mins | 0.43 (0.34, 0.54) |  | 0.60 (0.46, 0.80) |  |
| **In-hospital mortality or discharge to hospice** |  | <.0001 |  | 0.0289 |
| DIDO ≤ 90mins | Ref. |  |  |  |
| DIDO 91-180 mins | 0.78 (0.66, 0.92) |  | 0.80 (0.64, 1.01) |  |
| DIDO 181-270 mins | 0.66 (0.55, 0.80) |  | 0.81 (0.63, 1.04) |  |
| DIDO >270 mins | 0.49 (0.40, 0.59) |  | 0.67 (0.52, 0.87) |  |

Models adjusted for age, sex, insurance status, atrial fibrillation/flutter, prior stroke/TIA, Coronary Artery Disease / Myocardial Infarction, heart failure, carotid stenosis, diabetes, peripheral vascular disease, hypertension, dyslipidemia, prosthetic heart valve, smoker, renal insufficiency, NIHSS score, antiplatelet or anticoagulant, systolic blood pressure, INR, US region, rural location, teaching hospital, number of beds, annual hemorrhagic stroke volume, stroke center status.
<sup>†</sup>Beta Regression using transformed scores have been fit
Discharge mRS is reported among those with available data N=5,836
Discharge ambulatory events were modeled among patients independently ambulatory prior to admission N=5,961

Figures 1a and 1b depict the temporal trends of overall increasing DIDO times from 2019-2022 for ICH and SAH patients, respectively. Median DIDO times for patients with ICH increased from 143.5 min in the first quarter of 2019 to to 179.5 min in the third quarter of 2022, and increased from 164.0 min to 193.0 min for patients with SAH.

**Figure 1a.**
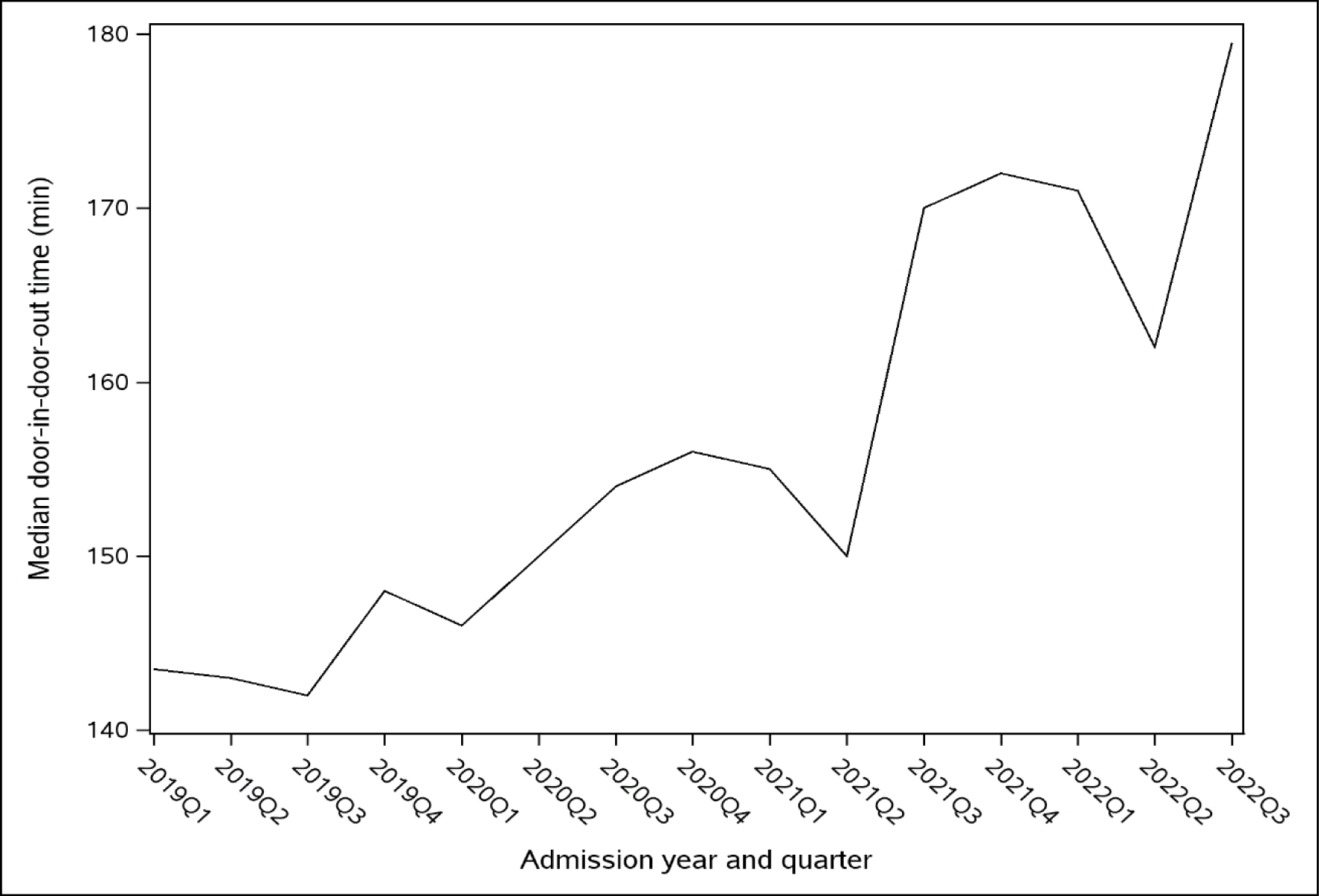
Temporal trends of door-in-door-out time by referring admission quarters for intracerebral hemorrhage patients

**Figure 1b.**
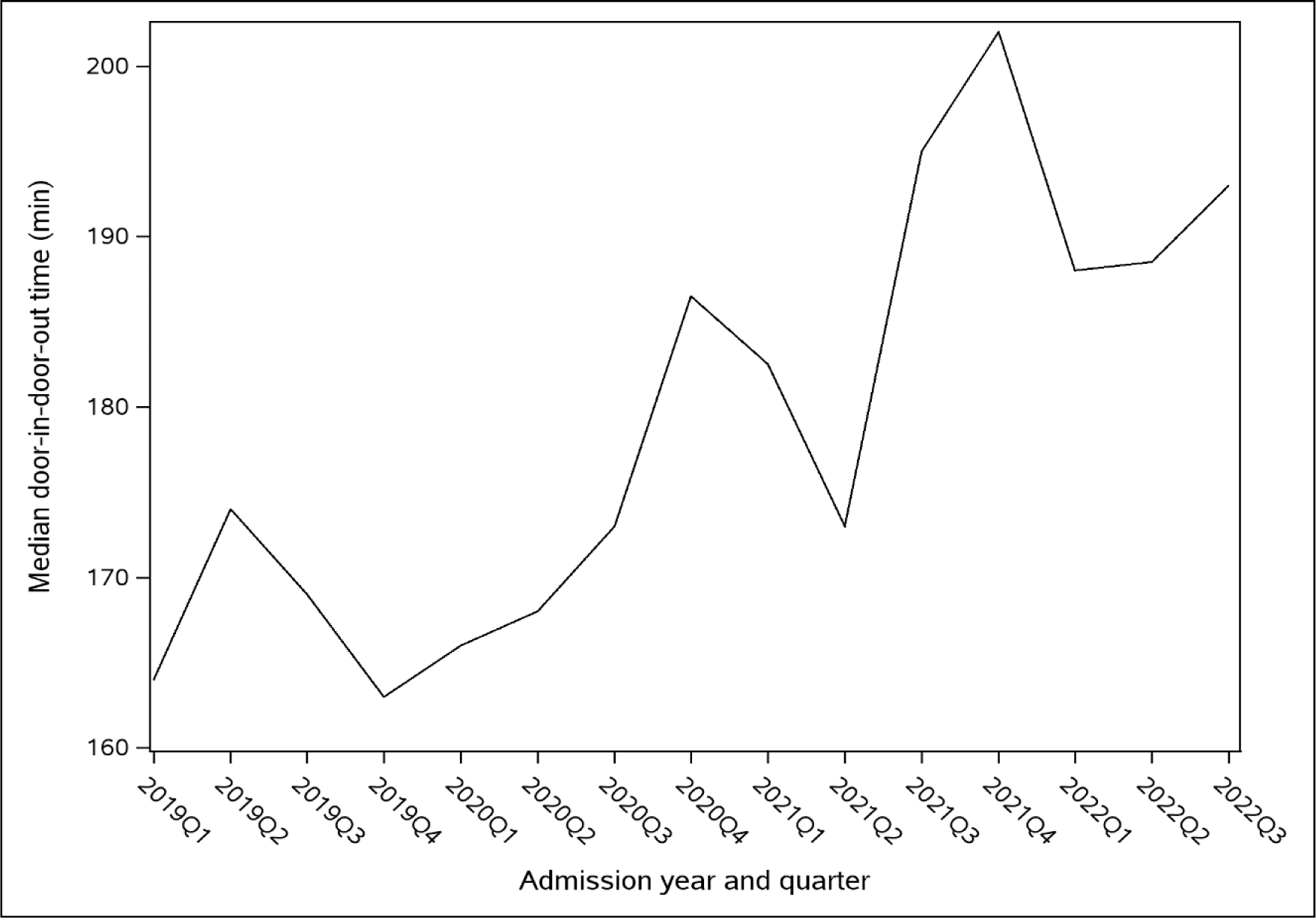
Temporal trends of door-in-door-out time by referring admission quarters for subarachnoid hemorrhage patients

## DISCUSSION

In this national study of interhospital transfers for hemorrhagic stroke, DIDO time was associated with higher odds of good functional outcome (mRS 0-3 vs 4-6) at discharge in the unadjusted analyses for both ICH and SAH patients, but these associations became statistically non-significant after adjustment for demographic factors, medical history, admission characteristics (including stroke severity) and receiving-hospital characteristics. Additionally, increasing DIDO times were associated with lower odds of in-hospital mortality and ability to ambulate independently at discharge for patients with ICH and SAH. After adjustment for covariates, the effect sizes of these associations were reduced, but the results remained statistically significant. These results were contradictory to prior literature^16,17^ and to our hypothesis that shorter DIDO times would be associated with more favorable outcomes.

While the results from our study are surprising, they do align with the findings from a recent secondary analysis of the RACECAT randomized clinical trial examining the effect of bypassing the closest stroke center in patients with ICH.^24^ In this secondary analysis of the multicenter, cluster-randomized trial conducted from 2017-2020, the authors found that direct transfer to an EVT-capable stroke center vs the nearest local stroke center resulted in worse functional outcomes at 90 days in patients with ICH.^24^ One potential explanation for these findings in RACECAT was patients with ICH transferred to EVT-capable stroke centers may have had delayed emergency stabilization and ICH “bundle of care” measures as compared to patients directly transported to local stroke centers. Results from INTERACT3^7^ and other literature^25^ have recently demonstrated that early bundled care for ICH, including antithrombotic reversal,^8^ appropriate management of blood pressure, hyperglycemia and pyrexia led to improved functional outcomes.^7^ While we were unable to measure the presence or absence of ICH care bundles in the current study, one interpretation of our findings is that longer DIDO times at transferring facilities may have afforded more adequate time for emergency stabilization measures, which could have translated into better outcomes from ICH and SAH.

However, it is important to consider the possibility of residual confounding that we were not able to account for. Stroke severity was approximated using NIHSS.^26^ A previous validation study showed that NIHSS was superior to GCS to identify patients at risk of poor early functional outcomes and large hematoma volumes.^27^ However, the possibility exists that other measures of severity (e.g. ICH Score, Hunt Hess) would more precisely capture the status of patients with hemorrhagic stroke. These additional stroke severity measures were not able to be included due to missingness. Due to this discrepancy, patients may have been less likely to be triaged to a comprehensive stroke center by EMS, or fail to screen-in on triage stroke scales in the waiting room of the emergency department. Finally, there is a possibility that baseline NIHSS at the initial hospital is less predictive than subsequent NIHSS scales obtained at the receiving hospital.

Our results show that DIDO times increased for patients with hemorrhagic stroke from 2019 to 2022. The study period in this analysis is largely contained within the COVID pandemic, which was associated with delays in stroke care^28^ and higher in-hospital mortality.^29^ Increasing DIDO times could also reflect concerning trends in hospital and ED overcrowding^30^ and delays in finding available neurological ICU hospital beds or dedicated inpatient stroke units, which previous studies have associated with worse outcomes.^16,17^

While the results from the current study represent a benchmark for current management of hemorrhagic stroke, the landscape of care may be changing. Improvements in functional outcomes and reduced mortality were recently demonstrated in the ENRICH trial of ICH evacuation using early, minimally invasive parafascicular surgery.^31,32^ Recent positive trials in both the medical and surgical acute management of ICH may herald a paradigm shift in the approach to hemorrhagic stroke management, and the results of the current study will need to be updated as the standard of care evolves in the coming years.

### Limitations

This study has several limitations. First, the analysis is limited to data that was entered by the *receiving* hospital about the *transferring* hospital encounter. This includes the primary predictor variable of DIDO time. This could skew cases to shorter transfer times and sicker patients; however, the median DIDO time recorded in this sample was 155 (101.0-243.0) minutes for ICH and 179.0 (120.0-273.0) minutes for SAH, while a previous study of DIDO time based solely on transferring hospital data was 178 minutes for all hemorrhagic strokes, which suggests these data are consistent with previous analyses of this population.^15^ Data from receiving hospitals had high rates of missingness for DIDO time. Thus, demographic and clinical characteristics of patients with missing DIDO time and DIDO >24 hours were compared with the study sample. Differences were observed in the SAH group which had higher proportions of mRS of 5 or 6 in the patients that were missing DIDO times (Supplemental Table 3 and 4), but were otherwise similar.

Second, ordinal measures of mRS were not able to be examined, as they failed to meet the proportional odds assumption. It is possible there is a subset of patients that may have a different relationship between DIDO time and discharge mRS that is not able to be elucidated in the current dichotomous analysis of discharge mRS.

### Conclusions

In this retrospective cohort study using a large national quality database, door-in-door-out times were inversely related to in-hospital mortality, and ability to ambulate independently at discharge, but not discharge mRS for patients with hemorrhagic stroke. These findings may suggest that a longer period of stabilization in the initial ED may be associated with better outcomes from hemorrhagic stroke and that current interhospital transfer protocols are currently designed to expedite transfer of the sickest patients. Prospective studies are needed to determine whether early or delayed transport with ED stabilization is optimal for care of patients with hemorrhagic stroke.

## Data Availability

Data is available through an application to the American Heart Association

## Acknowledgements

IA and BA had full access to all the data in the study and take responsibility for the integrity of the data and accuracy of the data analysis.

## Funding

The authors received no external sources of funding for this study. RR receives funding from the National Institute of Health though this work was not supported by this grant. PP receives funding from the National Institutes of Health Agency for Healthcare Research and Quality (AHRQ) (K08HS029208). SP receives funding from the AHRQ (R18HS027264) and National Institute of Neurological Disorders and Stroke (NINDS) (U24NS107233, R25NS125609; U01NS131797). KNS reported grants from the National Institutes of Health, American Heart Association, Hyperfine, Biogen, and Bard; serves on the data safety monitoring board for Zoll and Sense; serves on the scientific advisory board for CSL Behring, Astrocyte, and Rhaeos; and holds equity in Alva outside the submitted work; in addition, Dr Sheth had a patent for Alva issued. BM reported grants from the National Institutes of Health, American Heart Association, and Duke Office of Physician-Scientist Development outside the submitted work. WJM receives funding from NIH and PCORI, however this work was not supported by any of these grants.

To the best of our knowledge, the remaining authors have no conflict of interest, financial or otherwise. The Get With The Guidelines®–Stroke (GWTG-Stroke) program is provided by the American Heart Association. GWTG-Stroke is sponsored, in part, by Novartis, Novo Nordisk, AstraZeneca, Bayer and HCA Healthcare. This project was supported by the Hemorrhagic Stroke Data Challenge.

## Role of the Funder

There were no external funders of this study, however The American Heart Association provided input in design of the study, collection, analysis, review, and approval of the manuscript for publication.

## Notes

### Competing Interest Statement

RR receives funding from the National Institute of Minority Health and Health Disparities though this work was not supported by this grant. PP receives funding from the National Institutes of Health Agency for Healthcare Research and Quality (AHRQ) (K08HS029208). SP receives funding from the AHRQ (R18HS027264) and National Institute of Neurological Disorders and Stroke (NINDS) (U24NS107233, R25NS125609; U01NS131797). KNS reported grants from the National Institutes of Health, American Heart Association, Hyperfine, Biogen, and Bard; serves on the data safety monitoring board for Zoll and Sense; serves on the scientific advisory board for CSL Behring, Astrocyte, and Rhaeos; and holds equity in Alva outside the submitted work; in addition, Dr Sheth had a patent for Alva issued. BM reported grants from the National Institutes of Health, American Heart Association, and Duke Office of Physician-Scientist Development outside the submitted work. WJM receives funding from NIH and PCORI, however this work was not supported by any of these grants. To the best of our knowledge, the remaining authors have no conflict of interest, financial or otherwise. The Get With The Guidelines®–Stroke (GWTG-Stroke) program is provided by the American Heart Association. GWTG-Stroke is sponsored, in part, by Novartis, Novo Nordisk, AstraZeneca, Bayer and HCA Healthcare. This project was supported by the Hemorrhagic Stroke Data Challenge.

### Funding Statement

The authors received no external sources of funding for this study. The Get With The Guidelines®–Stroke (GWTG-Stroke) program is provided by the American Heart Association. GWTG-Stroke is sponsored, in part, by Novartis, Novo Nordisk, AstraZeneca, Bayer and HCA Healthcare. This project was supported by the Hemorrhagic Stroke Data Challenge.

### Author Declarations

Each participating hospital received either human research approval to enroll cases without individual patient consent under the common rule, or a waiver of authorization and exemption from subsequent review by their institutional review board. The Duke Clinical Research Institute serves as the data analysis center and has an agreement to analyze the aggregate limited data for research purposes. The Institutional Review Board at Duke University Health approved this study. IQVIA (Parsippany, New Jersey) serves as the data collection and coordination center.

